# Epidemiology and Survival outcomes of HIV-associated cervical cancer in Nigeria

**DOI:** 10.1101/2023.08.08.23293820

**Authors:** Jonah Musa, Masha Kocherginsky, Francis A. Magaji, Ali J. Maryam, Joyce Asufi, Danjuma Nenrot, Kristen Burdett, Neelima Katam, Elizabeth N. Christian, Nisha Palanisamy, Olukemi Odukoya, Olugbenga A. Silas, Fatimah Abdulkareem, Philip Akpa, Kabir Badmos, Godwin E. Imade, Alani S. Akanmu, Demirkan B. Gursel, Yinan Zheng, Brian T. Joyce, Chad J. Achenbach, Atiene S. Sagay, Rose Anorlu, Jian-Jun Wei, Folasade Ogunsola, Robert L. Murphy, Lifang Hou, Melissa A. Simon

## Abstract

**Introduction:** Invasive cervical cancer (ICC) is an HIV-associated cancer that is preventable and precancerous stages including early ICC stages could be detected through screening offering opportunities for treatment and cure. The high incidence in women living with HIV and late presentation often at advanced stages of ICC with limited treatment facilities often result in early mortality. We sought to compare the epidemiologic characteristics and survival differences in HIV status of ICC patients in Nigeria.

**Methods:** We conducted a cohort study at two federal academic hospital-based research sites in Jos University Teaching Hospital, and Lagos University Teaching Hospital Nigeria, between March 2018 and September 2022. We enrolled women with histologically confirmed ICC with known HIV status, and FIGO staging as part of the United States of America’s National Institutes of Health/National Cancer Institute funded project titled ‘Epigenomic Biomarkers of HIV-Associated Cancers in Nigeria’. The primary outcome was all-cause mortality with assessment of overall survival (OS) and time to death after ICC diagnosis. OS distribution was estimated using the method of Kaplan-Meier and compared between groups using the log-rank test.

**Results:** A total of 239 women with confirmed ICC were enrolled and included in this analysis, of whom 192 (80.3%) were HIV-negative (HIV-/ICC+), and 47 (19.7%) were HIV-positive (HIV+/ICC+). The HIV+/ICC) patients were younger with median age 46 (IQR: 40-51) years compared to 57 (IQR: 45-66) among HIV-/ICC+) (P<0.001. Squamous cell carcinoma was the commonest histopathologic variant in 80.4% of ICC diagnosis, moderately differentiated tumor grade in 68.1% in both groups. HIV+/ICC+ diagnosis was at FIGO advanced stages in 64.9% compared to 47.9% in HIV-/ICC+. The HIV-/ICC+ women had better OS compared to HIV+/ICC+ participants (p=0.018), with 12-month OS 84.1% (95%CI: 75% - 90%) and 67.6% (95%CI: 42%-84%) respectively.

**Conclusion:** ICC is diagnosed at a relatively young age in women living with HIV, with a significantly lower overall survival probability compared to women without HIV. The trend of presentation and diagnosis at advanced stages in women living with HIV could partly explain the differences in overall survival.

## Introduction

The World Health Organization (WHO) global estimates showed that 604,127 new cases of invasive cervical cancer (ICC) and 341,831 deaths occurred in 2020.(1) It is the fourth-most common cancer among women worldwide, the leading cause of cancer-related death in women in Africa and is ranked in the top three cancers affecting women younger than 45 years. (2) ICC is the second-most diagnosed cancer in West Africa and the fourth-most common in Nigeria(3) with a high annual incidence and mortality.(4) The 2020 WHO estimates showed a rising burden and mortality in countries with low human development index compared to a declining incidence and mortality in high human development index countries.(1) The high burden of human immunodeficiency (HIV) virus infection has been shown to be a significant aggravating factor with ICC incidence and this has been confirmed in a prior systematic analysis showing that women living with HIV had at least 6 times greater risk of developing ICC compared to women without HIV. (5)

Late presentation and diagnosis, lack of access to care, low resources, and ineffective treatment of ICC have contributed to the high mortality and poor survival for cases of ICC in Nigeria and most low- and middle-income countries (LMICs). (6–8) The delay in diagnosis and related poor prognosis has necessitated the efforts towards cervical cancer prevention through human papillomavirus (HPV) vaccination programs and early detection and treatment of precancerous stages of the cervix, particularly in high-risk groups. This is particularly important in sub-Saharan Africa contributing to over 24.5% of global mortality with fewer national HPV vaccination programs.(4) Previous research from Jos, Nigeria showed that over 72% of women with cervical cancer were diagnosed at advanced clinical stages, with survival of only 20% after 2 years post-diagnosis. (9) These findings further support the WHO initiative on HPV vaccination, high-performing screening strategies, and effective treatments for precancerous conditions including early stage ICC. (10)

Previous epidemiologic studies have focused on the survival outcomes of patients with cervical cancer based on stage at diagnosis, access to treatment by surgery or chemoradiation, but few have compared survival outcomes by HIV status.(8, 9, 11, 12) Furthermore, studies on survival outcomes by HIV status have reported inconsistent effects on cervical cancer mortality.(12–15) This manuscript provides the descriptive epidemiologic characteristics of women diagnosed with invasive cervical cancer enrolled at 2 large tertiary academic medical centers in Nigeria, adding to previous survival data reported(9), expanding upon survival differences by HIV status.

## Materials and Methods

### Study Design and Setting

We conducted a cohort study at two academic hospital-based research sites in Jos (Jos University Teaching Hospital, UniJos) and Lagos (Lagos University Teaching Hospital, UniLag), Nigeria. We enrolled women with histologically confirmed invasive cervical cancer, with and without HIV, as part of the United States of America’s National Institutes of Health/National Cancer Institute funded project titled ‘Epigenomic Biomarkers of HIV-Associated Cancers in Nigeria’ (U54CA221205) (see inclusion below).

### Study participants and data collection

Eligible study participants were approached for enrollment at the two research sites between March 2018 and September 2022. Trained research staff screened and enrolled women aged 18 or older who were not pregnant, no history of hysterectomy, and were not undergoing cancer treatment at the time of recruitment. Participants were classified into two cohorts based on histopathological confirmation of ICC and HIV diagnosis: 1) HIV-positive women with ICC (study group: HIV+/ICC+), 2) HIV-negative women with ICC (control group): HIV-/ICC+). Participants’ clinical and demographic information were obtained by interviewer-administered questionnaires at the time of enrollment into the study. The intent and procedures of the study were disclosed to the study participants and informed consent was obtained. The study protocol was reviewed and approved by the Institutional Review Boards (IRB) at UniJos (JUTH/DCS/ADM/127/XXVII/630) and UniLag (CMUL/HREC/02/22/327/V4) in Nigeria, and Northwestern University (NU) in the USA (STU00207051).

### HIV diagnosis and care information

The HIV status of study participants receiving care and treatment at the Presidential Emergency Plan for AIDS Relief (PEPFAR) program of the two participating institutions was extracted from the adult HIV treatment and care database. Women with unknown HIV status had rapid HIV diagnostic testing according to the national HIV testing serial algorithm, which involved the use of the Rapid Determine Test, Unigold, and STAT Pack rapid HIV diagnostic test kits. All HIV-infected women receiving care in the PEPFAR program at both study sites were on antiretroviral therapy (ART) at the time of study enrollment. Those whose HIV infection was diagnosed at enrollment were provided appropriate HIV counseling and linked to care and commencement of ART in the PEPFAR program of the participating institutions in accordance with the Nigerian National ART Guidelines(16).

### Clinical and histopathologic diagnosis of Invasive cervical cancer

Suspected ICC cases were evaluated by the gynecologic oncology team at the Jos University Teaching Hospital (UniJos) and Lagos University Teaching Hospital (UniLag). Diagnostic evaluation of ICC included examination under anesthesia (EUA), and clinical staging according to the international federation of gynecology and obstetrics (FIGO 2009). (17) During the EUA, cervical tumor tissue biopsy was obtained for histopathological diagnosis and tumor grading. Trained pathologists at each of the enrollment sites reviewed the biopsied tissues and provided histopathological diagnosis and tumor grading as: well-differentiated (grade 1), moderately differentiated (grade 2), and poorly differentiated (grade 3). The histopathology slides and paraffin-fixed blocks were shared with the Northwestern University Cancer Center Pathology Core team for verification. We also utilized the telepathology scanners established between the 2 primary sites and the Northwestern Telepathology team(18) for quality assurance of tissue diagnosis of all participants enrolled in the study. Specifically, additional H&E staining was performed and evaluated at the Pathology Core Facility of Northwestern University and was discussed by telepathology for all sites (see supplementary file).

### Care and treatment of patients with invasive cervical cancer

As previously reported(9), the paucity of trained gynecologic oncology specialist in Nigeria, and limited access to standard chemoradiation centers(9, 19), made it difficult to provide a uniform standard of care for patients with invasive cervical cancer during the study period. However, patients diagnosed at early stages of the disease (FIGO stages 1-IIA) received modified radical hysterectomy followed by adjuvant chemotherapy alone or concomitant with radiotherapy either onsite or by referral to functional radiotherapy centers within Nigeria. Those with advanced stage disease received either symptomatic care (including correction of anemia, treatment of infections, and pain control) or referral for chemotherapy or primary chemoradiation. Follow up data were obtained from the gynecology outpatient clinic records, radio-oncology clinics records, and via phone calls (patient’s and next-of-kin phone contacts were obtained at diagnosis) to ascertain and update treatment status.

### Outcome and ascertainment of death

The primary clinical outcome analyzed in this study was all-cause mortality with assessment of overall survival and time to death after invasive cervical cancer diagnosis. Overall survival (OS) is defined as time from ICC diagnosis to death with censoring at the date of last available follow up as obtained from clinic visits or telephone contact up to September 30^th^, 2022. The use of telephone for cancer follow up has been documented in a prior study in Jos Nigeria and in other studies in Africa.(9, 20)

### Data management and statistical analysis

Study data were collected and managed using REDCap electronic data capture tools(21, 22) with custom databases developed by the Biomedical Informatics and Statistics Core (BISC). REDCap (Research Electronic Data Capture) is a secure, web-based software platform designed to support data capture for research studies, providing 1) an intuitive interface for validated data capture; 2) audit trails for tracking data manipulation and export procedures; 3) automated export procedures for seamless data downloads to common statistical packages; and 4) procedures for data integration and interoperability with external sources. Data entry at both UniJos and UniLag was done by the site data managers team using REDCap(21) with periodic synchronization of the data with the Northwestern University REDCap server. Data quality reports were generated using R(23) and R Markdown by the NU BISC team. Details of our experience with data management of this project on REDCap have been published elsewhere.(24)

### Statistical analysis

Descriptive statistics were used to summarize baseline subject characteristics for ICC+/HIV+ and ICC+/HIV-groups, and overall. Continuous variables were summarized using median and interquartile range (IQR) and compared between groups using the Wilcoxon rank sum test. For categorical variables, frequencies and percentages were reported, and proportions were compared using Fisher’s exact test. Frequencies of missing observations were tabulated, and missing observations were not included in these descriptive analyses.

Primary objective was to determine the association between HIV status and overall survival (OS) among ICC+ patients. OS distribution was estimated using the method of Kaplan-Meier, and survival estimates are reported with the corresponding 95% confidence bands, 25^th^, median, and 75^th^ percentiles survival times are presented. Overall survival was compared between groups using the log-rank test. Overall survival was estimated for HIV+ and HIV-groups, and by study site.

Statistical analyses were performed using R v. 4.2.2 statistical software(23), and using gtsummary(25), survival(26) and survminer(27) packages.

## Results

Between March 18^th^, 2018, and September 30^th^, 2022, a total of 239 women with confirmed invasive cervical cancer were enrolled and are included in this analysis, of whom 192 (80.3%) were HIV-negative (HIV-/ICC+), and 47 (19.7%) were HIV-positive (HIV+/ICC+).

The median age at study enrollment was 53 years (IQR: 44-65) among the ICC+ participants overall. HIV-positive patients were younger with median age 46 (IQR: 40-51) years compared to 57 (IQR: 45-66) among HIV-negative (P-value<0.001); had a higher level of education with only 10.6% having no formal education as compared to 32.6% in the HIV-groups (p<.001); and a higher number of lifetime sexual partners with median of 3 (IQR: 1-4) partners among HIV+ patients vs. 1 (IQR: 1-2) in the HIV-group (p<0.001). Women without HIV had a higher number of childbirths (76.5% vs. 58.7% had 4 or more live births among HIV- and HIV+ women, respectively; p<0.001) but comparable age at first childbirth (median 20 years old in both groups) and BMI (median 25 in both groups) to women with HIV. The most common histopathologic variant of ICC was squamous cell carcinoma in 80.4% of all cases and was similar among HIV+ and HIV-women (79.2% and 80.6%, respectively). The majority of patients (68.1%) had moderately differentiated tumor grade (61.5% and 69.1% among HIV+ and HIV-patients, respectively), and the histopathologic variants and tumor grading were comparable in both groups. HIV+ women tended to be diagnosed with advanced stage more frequently (64.9%) than HIV-women (47.9%) although the difference was not statistically significant (p=0.10). Other descriptive statistics of the participants with ICC by HIV status have been summarized in Table 1.

**Table 1.**
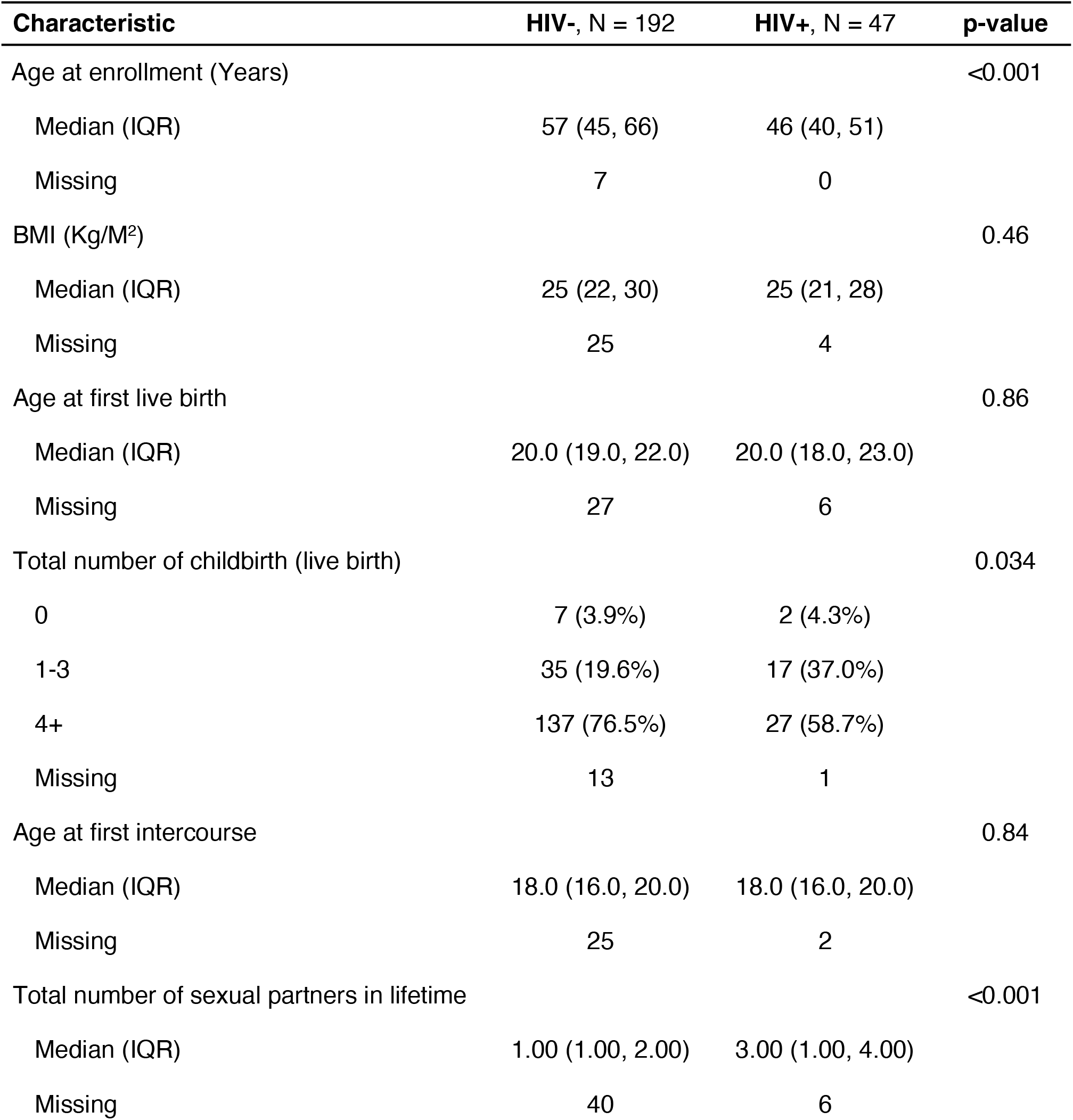
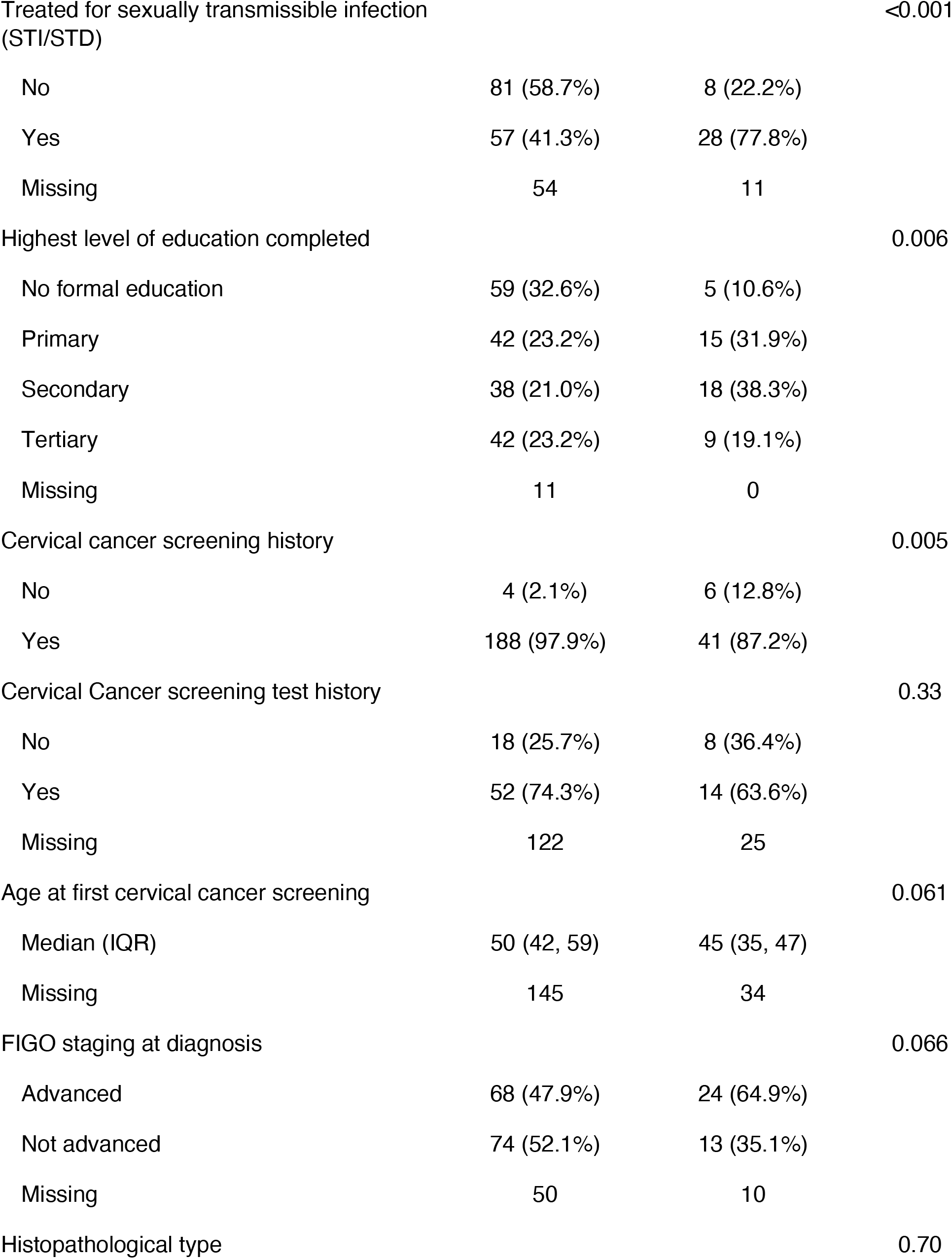
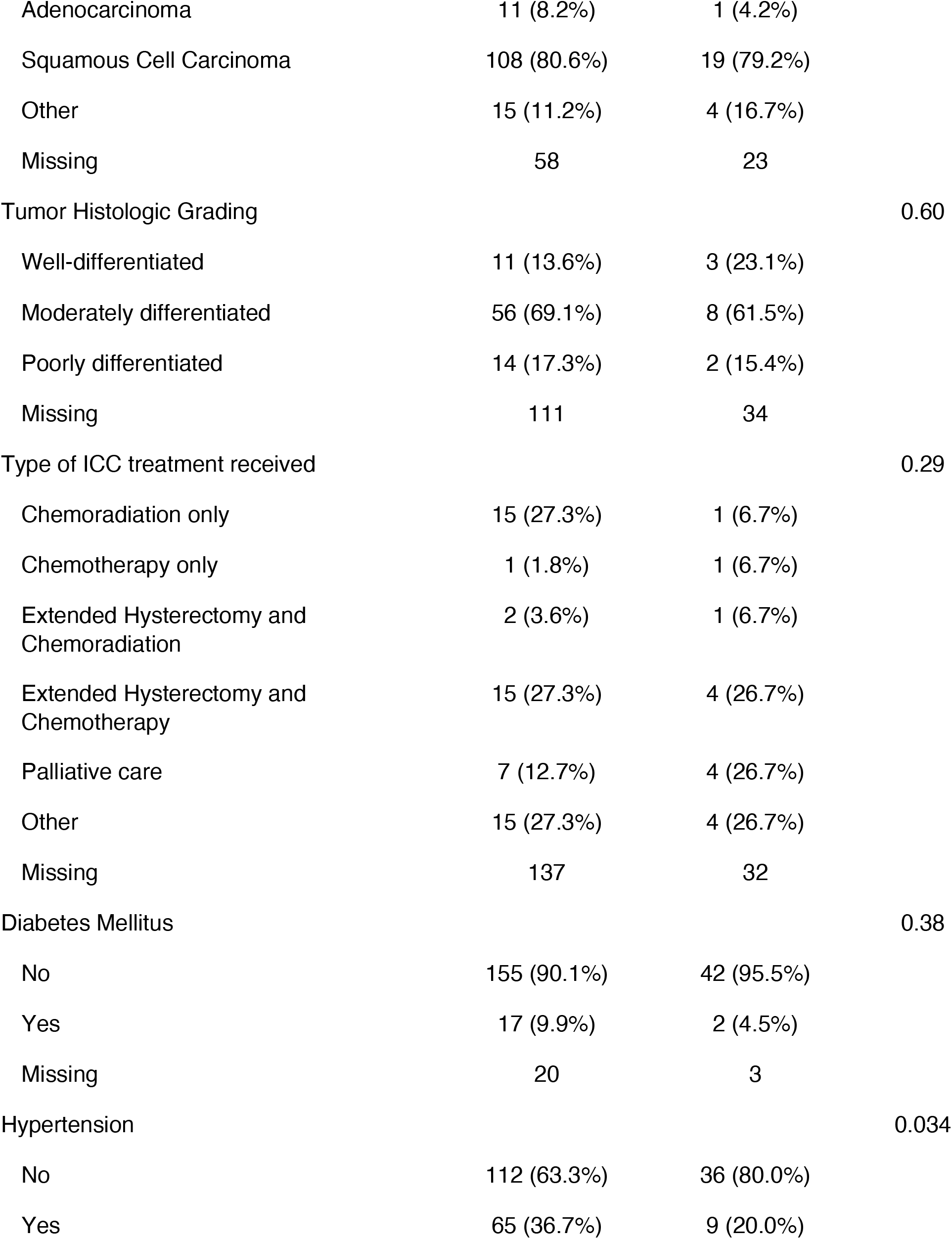
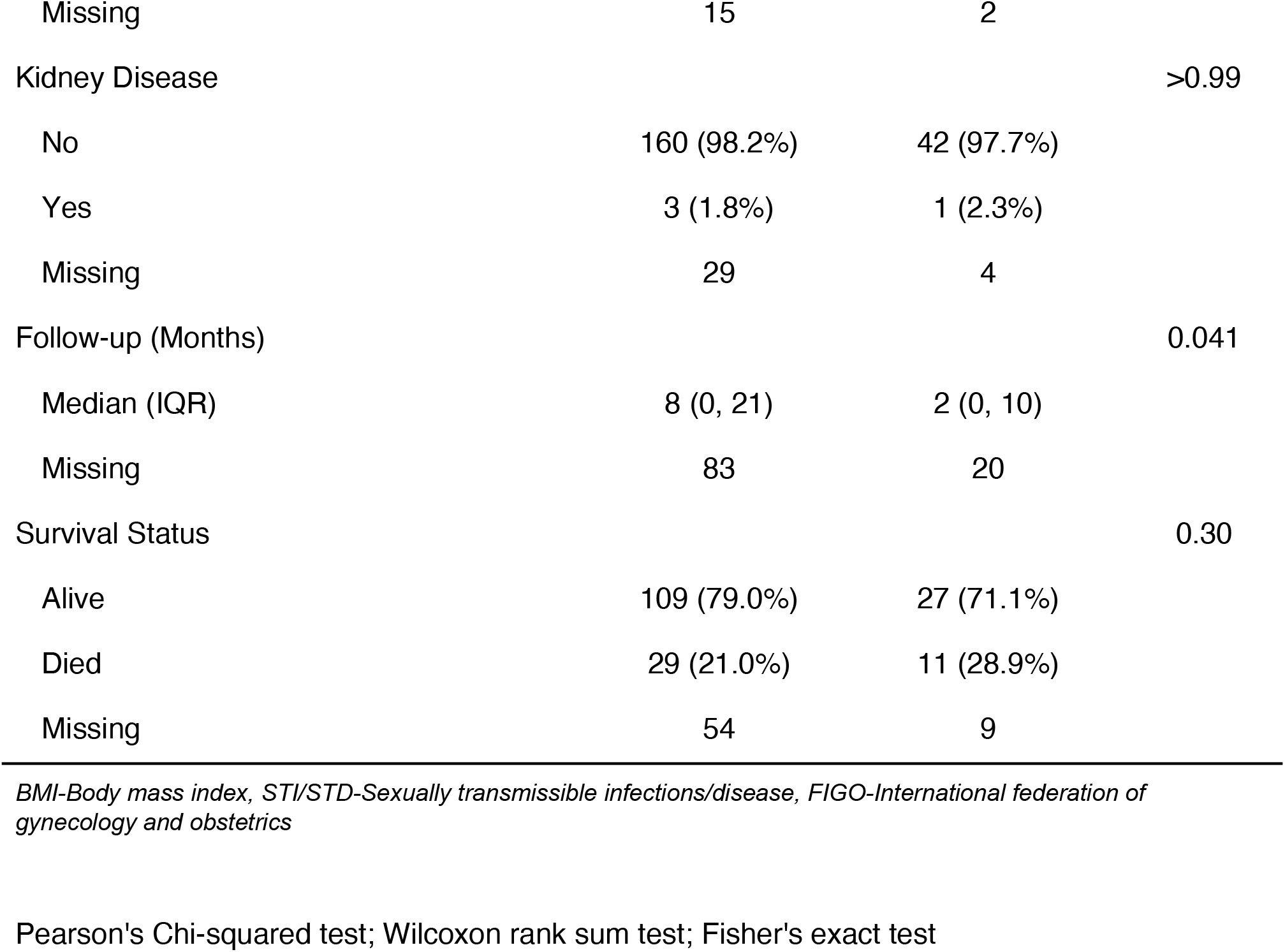
Baseline socio-demographic characteristics of the 239 study participants diagnosed with invasive cervical cancer by HIV status (JUTH and LUTH combined)

| Characteristic | HIV-, N = 192 | HIV+, N = 47 | p-value |
| --- | --- | --- | --- |
| Age at enrollment (Years) |  |  | <0.001 |
| Median (IQR) | 57 (45, 66) | 46 (40, 51) |  |
| Missing | 7 | 0 |  |
| BMI (Kg/M<sup>2</sup>) |  |  | 0.46 |
| Median (IQR) | 25 (22, 30) | 25 (21, 28) |  |
| Missing | 25 | 4 |  |
| Age at first live birth |  |  | 0.86 |
| Median (IQR) | 20.0 (19.0, 22.0) | 20.0 (18.0, 23.0) |  |
| Missing | 27 | 6 |  |
| Total number of childbirth (live birth) |  |  | 0.034 |
| 0 | 7 (3.9%) | 2 (4.3%) |  |
| 1-3 | 35 (19.6%) | 17 (37.0%) |  |
| 4+ | 137 (76.5%) | 27 (58.7%) |  |
| Missing | 13 | 1 |  |
| Age at first intercourse |  |  | 0.84 |
| Median (IQR) | 18.0 (16.0, 20.0) | 18.0 (16.0, 20.0) |  |
| Missing | 25 | 2 |  |
| Total number of sexual partners in lifetime |  |  | <0.001 |
| Median (IQR) | 1.00 (1.00, 2.00) | 3.00 (1.00, 4.00) |  |
| Missing | 40 | 6 |  |
| Treated for sexually transmissible infection (STI/STD) |  |  | <0.001 |
| No | 81 (58.7%) | 8 (22.2%) |  |
| Yes | 57 (41.3%) | 28 (77.8%) |  |
| Missing | 54 | 11 |  |
| Highest level of education completed |  |  | 0.006 |
| No formal education | 59 (32.6%) | 5 (10.6%) |  |
| Primary | 42 (23.2%) | 15 (31.9%) |  |
| Secondary | 38 (21.0%) | 18 (38.3%) |  |
| Tertiary | 42 (23.2%) | 9 (19.1%) |  |
| Missing | 11 | 0 |  |
| Cervical cancer screening history |  |  | 0.005 |
| No | 4 (2.1%) | 6 (12.8%) |  |
| Yes | 188 (97.9%) | 41 (87.2%) |  |
| Cervical Cancer screening test history |  |  | 0.33 |
| No | 18 (25.7%) | 8 (36.4%) |  |
| Yes | 52 (74.3%) | 14 (63.6%) |  |
| Missing | 122 | 25 |  |
| Age at first cervical cancer screening |  |  | 0.061 |
| Median (IQR) | 50 (42, 59) | 45 (35, 47) |  |
| Missing | 145 | 34 |  |
| FIGO staging at diagnosis |  |  | 0.066 |
| Advanced | 68 (47.9%) | 24 (64.9%) |  |
| Not advanced | 74 (52.1%) | 13 (35.1%) |  |
| Missing | 50 | 10 |  |
| Histopathological type |  |  | 0.70 |

| **Characteristic** | **HIV-, N = 192** | **HIV+, N = 47** | **p-value** |
| --- | --- | --- | --- |
| Adenocarcinoma | 11 (8.2%) | 1 (4.2%) |  |
| Squamous Cell Carcinoma | 108 (80.6%) | 19 (79.2%) |  |
| Other | 15 (11.2%) | 4 (16.7%) |  |
| Missing | 58 | 23 |  |
| Tumor Histologic Grading |  |  | 0.60 |
| Well-differentiated | 11 (13.6%) | 3 (23.1%) |  |
| Moderately differentiated | 56 (69.1%) | 8 (61.5%) |  |
| Poorly differentiated | 14 (17.3%) | 2 (15.4%) |  |
| Missing | 111 | 34 |  |
| Type of ICC treatment received |  |  | 0.29 |
| Chemoradiation only | 15 (27.3%) | 1 (6.7%) |  |
| Chemotherapy only | 1 (1.8%) | 1 (6.7%) |  |
| Extended Hysterectomy and Chemoradiation | 2 (3.6%) | 1 (6.7%) |  |
| Extended Hysterectomy and Chemotherapy | 15 (27.3%) | 4 (26.7%) |  |
| Palliative care | 7 (12.7%) | 4 (26.7%) |  |
| Other | 15 (27.3%) | 4 (26.7%) |  |
| Missing | 137 | 32 |  |
| Diabetes Mellitus |  |  | 0.38 |
| No | 155 (90.1%) | 42 (95.5%) |  |
| Yes | 17 (9.9%) | 2 (4.5%) |  |
| Missing | 20 | 3 |  |
| Hypertension |  |  | 0.034 |
| No | 112 (63.3%) | 36 (80.0%) |  |
| Yes | 65 (36.7%) | 9 (20.0%) |  |
| Missing | 15 | 2 |  |
| Kidney Disease |  |  | >0.99 |
| No | 160 (98.2%) | 42 (97.7%) |  |
| Yes | 3 (1.8%) | 1 (2.3%) |  |
| Missing | 29 | 4 |  |
| Follow-up (Months) |  |  | 0.041 |
| Median (IQR) | 8 (0, 21) | 2 (0, 10) |  |
| Missing | 83 | 20 |  |
| Survival Status |  |  | 0.30 |
| Alive | 109 (79.0%) | 27 (71.1%) |  |
| Died | 29 (21.0%) | 11 (28.9%) |  |
| Missing | 54 | 9 |  |

*BMI-Body mass index, STI/STD-Sexually transmissible infections/disease, FIGO-International federation of gynecology and obstetrics*
Pearson's Chi-squared test; Wilcoxon rank sum test; Fisher's exact test

### Overall survival and differences in outcomes by HIV status

Following diagnosis and enrollment of participants with ICC, types of treatments received were not significantly different for women with HIV and without HIV as shown in Table 1. Follow-up for survival was available for 176 women (n=138 and n=38 in the HIV- and HIV+ groups, respectively), and 40 deaths were observed (n=29 and n=11 among HIV- and HIV+ patients, respectively). Median follow-up time among n=136 patients who were censored was 7.1 (IQR: 0-17.9) months. The overall survival outcomes of the entire ICC cohort are shown in the Kaplan-Meier curves in fig. 1. Median OS was not reached, and 12-month OS was 81.1 % (95%CI: 74.2% – 88.7%).

**Fig. 1:**
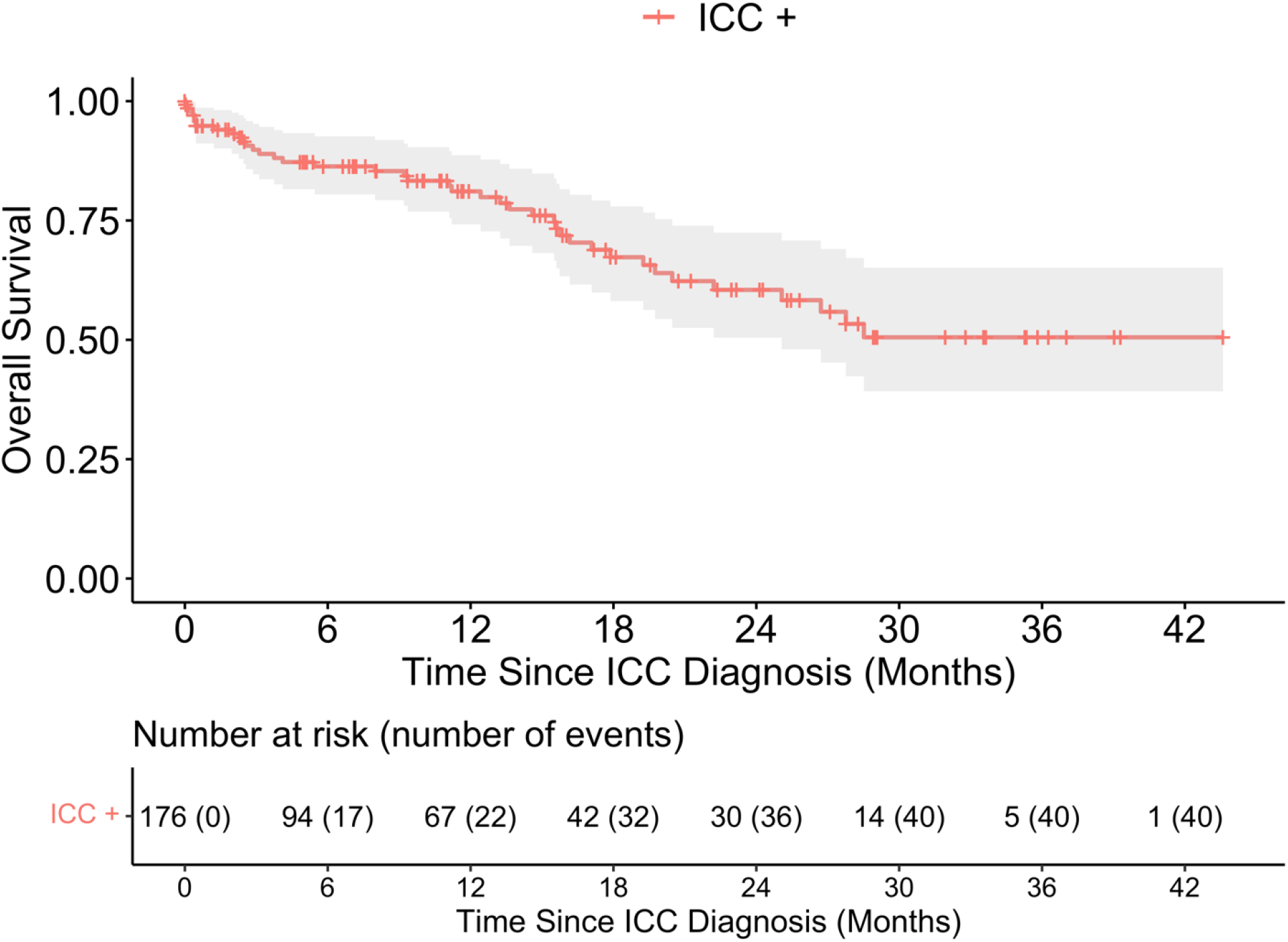
Overall survival of enrolled participants with invasive cervical cancer (ICC+). *Note: Of the 239 ICC+ participants, there were 40 events of mortality from 176 patients with available survival data.*

We also compared the survival outcomes by HIV status (Figure 2) and found that HIV negative women had better OS compared to participants who were HIV positive (p-=0.018), with 12-month OS 84.1% (95%CI: 75% - 90%) and 67.6% (95%CI: 42%-84%) among HIV- and HIV+ women, respectively. ICC participants diagnosed at early FIGO stage had better OS compared to participants at advanced stages of ICC (p-value=0.0045), with 94.0% (95%CI: 83% -98%) and 68.2% (95%CI: 53% -79%) among the early stage vs. advanced stage at diagnosis (Figure 3).

**Fig. 2:**
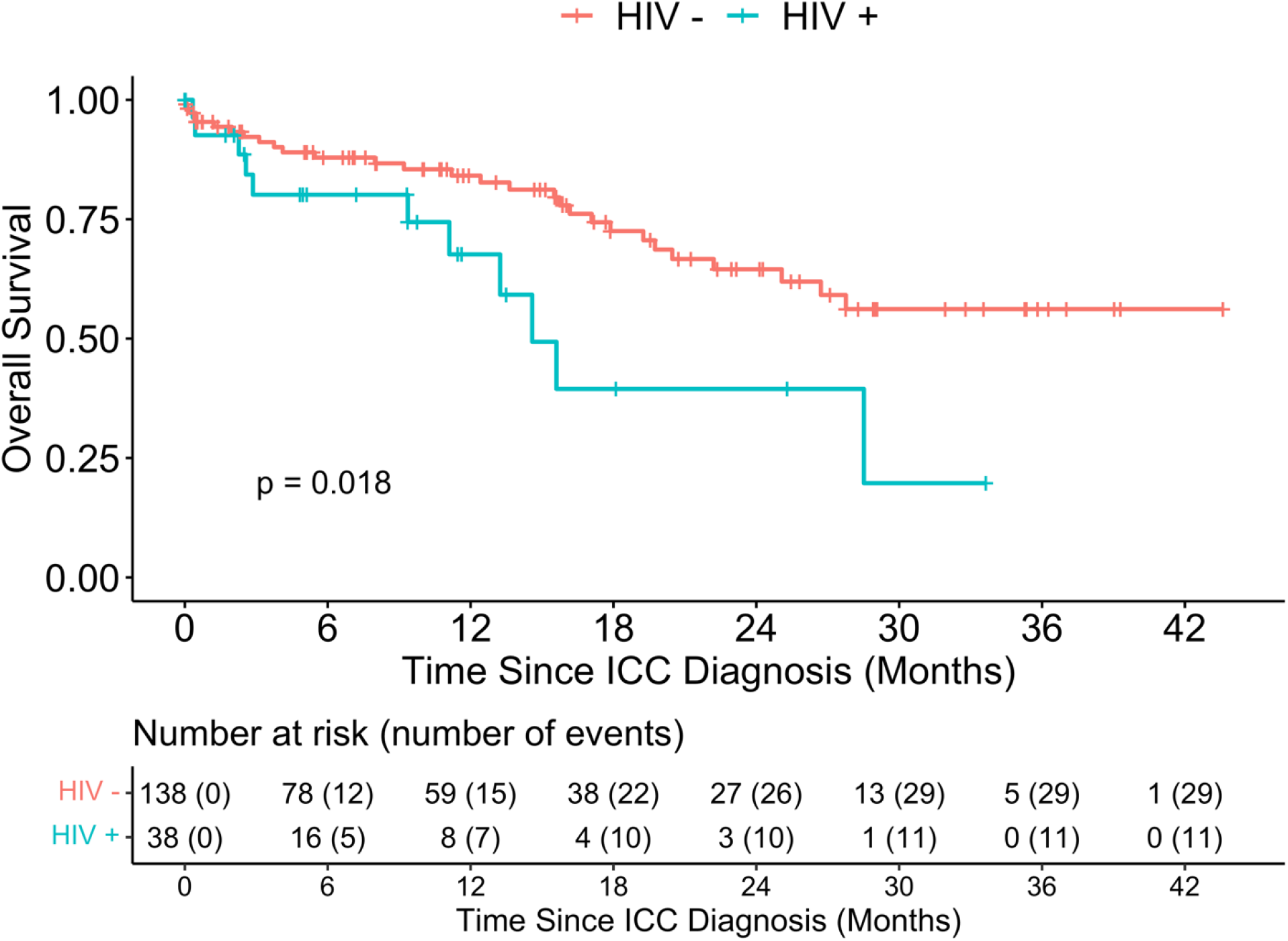
Overall survival distribution of patients with ICC stratified by HIV status.

**Fig. 3:**
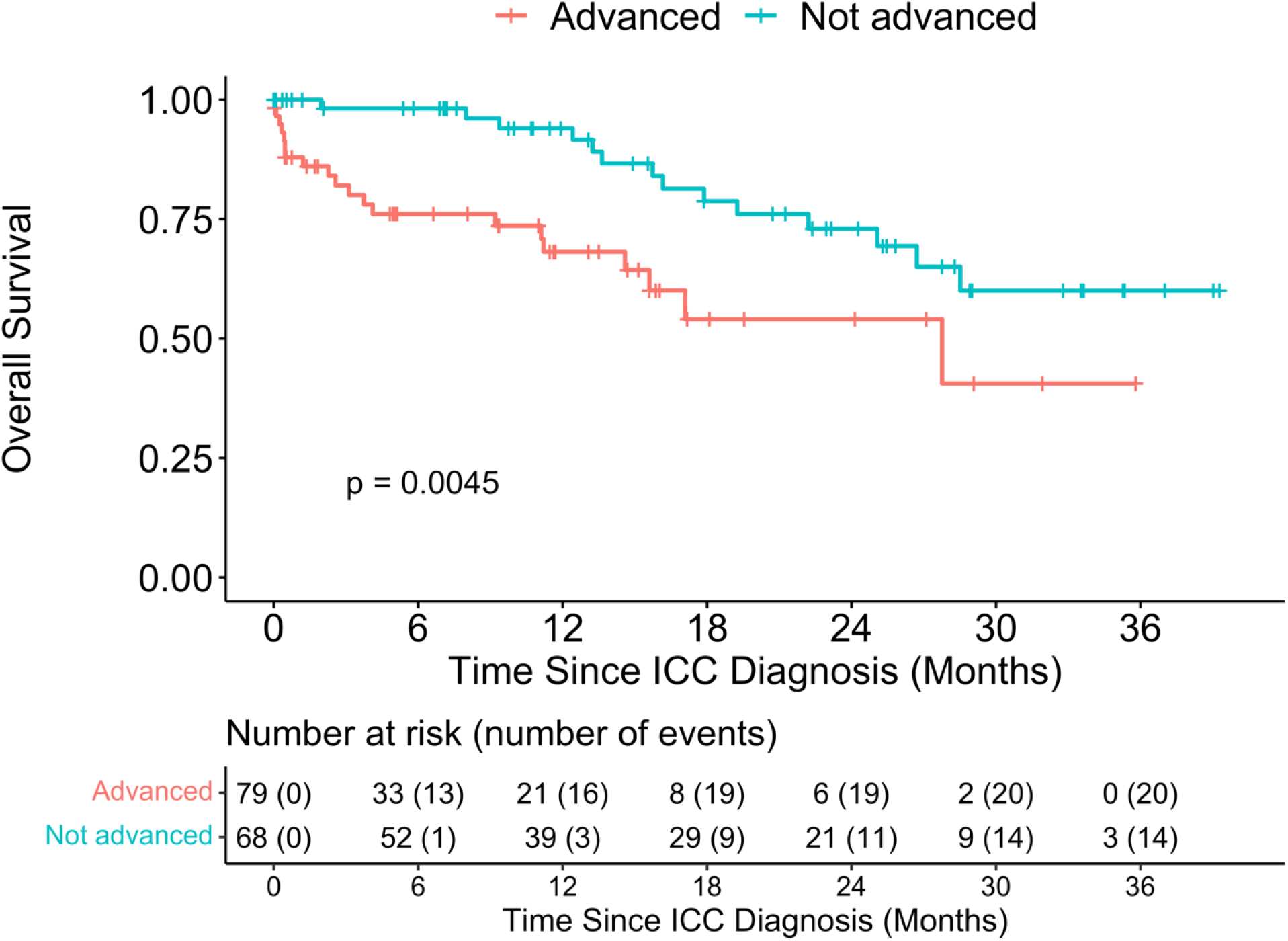
Overall survival distribution of patients with ICC stratified by FIGO stage at diagnosis (Advanced vs not Advanced) *Note: The FIGO stage at diagnosis was dichotomized as Advanced (stages IIB or more) versus Not Advanced or Early (stages IIA or less)*

## Discussion

This manuscript discusses the descriptive epidemiologic characteristics and survival outcomes of women with and without HIV diagnosed with invasive cervical cancer enrolled in two large tertiary academic medical centers in Nigeria. We found that women with HIV were 11 years younger at diagnosis of ICC compared to women without HIV. Although there were no significant differences in baseline BMI, FIGO staging at diagnosis, histopathologic variants, tumor grading and types of treatment received, we found that overall survival was significantly better in ICC participants without HIV compared to those living with HIV. Our findings were similar with a larger Zambian cohort of ICC patients that showed women with HIV were 10 years younger at diagnosis with a poorer prognosis compared to those without HIV.(14)

The relatively low survival rates in our primary analyses likely reflect the fact that treatment options for cervical cancer are greatly limited in most LMIC settings including Nigeria, with a paucity of trained gynecologic oncology specialists and limited cancer centers with chemoradiation facilities. A recent review has corroborated the poor treatment facilities, particularly the lack of radiotherapy centers to compensate the poor surgical services for gynecological cancers in sub-Saharan African settings.(19) These challenges have limited the use of and adherence to existing cervical cancer standard treatment guidelines in the management of patients diagnosed with ICC in Nigeria including the two primary enrollment sites in this report. Nevertheless, both enrollment sites have experienced gynecologists with expertise in surgical management of early-stage cervical cancer which was offered at diagnosis to patients with early-stage ICC. In addition, both sites had trained radiation oncologists onsite for chemotherapy and linkage to chemoradiation centers onsite or by referral.

The paucity of trained gynecologic oncologists and limited compliance to treatment guidelines is widespread in LMICs. This has been reported in a related study in Botswana in southern Africa that showed comparable findings of women with HIV being diagnosed with ICC at a much younger age compared to women without HIV, and the hazards of early mortality were significantly higher in women with HIV.(28) The effects of HIV on earlier ICC mortality are not clearly understood, but may be related to immune suppression with lower CD4 count.(28, 29) Further research in this population should explore this possibility.

The above findings have implications given the evidence of high incidence of cervical cancer in women living with HIV.(5) It is important to note that although previous reports have shown that women living with HIV were more likely to have a cervical cancer screening at younger ages(30), and our current data further add to the evidence that women living with HIV reported a younger age at first cervical cancer screening, the consistent reports showing that women living with HIV were diagnosed with ICC at a relatively younger age suggests that the transition between cervical precancer to ICC stages may be significantly shortened by HIV viremia. These findings should translate to public health policies that could build a stronger health system for preventing both HIV and human papillomavirus infections. Investing in primary prevention with human papillomavirus vaccination and strengthening cervical cancer screening for early detection of precancer, as well as offering effective treatment of precancerous lesions of the cervix in the general population (and especially in vulnerable populations such as women living with HIV). These strategies are effective and could make significant contributions to driving down the incidence of cervical cancer towards the WHO “elimination” target.(10)

Our data showed comparable histologic types of ICC and tumor grading irrespective of HIV status. As in most literature, the most common histologic variant found in both HIV positive, and HIV negative cervical cancer tumors is squamous cell carcinoma variants. A previous study report of a retrospective cervical cancer cohort found that women living with HIV were significantly more likely to be diagnosed at advanced stages of cancer compared to HIV negative women.(31) Our data showed a similar trend with 64.9% of HIV positive participants diagnosed at FIGO advanced stages compared to 47.9% in women who were HIV negative (P-value=0.066; Table 1). The significant factors predicting early mortality in the retrospective cohort of cervical cancer patients in Ethiopia were older age of patients, advanced stage of cancer diagnosis, and anemia.(31)

A previous histopathologic study of ICC showed that the commonest variant in both HIV positive and negative patients is the squamous cell type, and as in our study cohort is diagnosed at younger age in HIV-infected patients compared to HIV-negative patients.(32) The study also reported that HIV-infected patients were diagnosed at advanced FIGO stages of the disease and that the adenocarcinoma variant was commoner in HIV-positive patients.(32) The prognostic significance of adenocarcinoma in HIV infected women is not well understood but studies to understand cancer histology and differences in age at ICC diagnoses showed that adenocarcinoma types are systematically diagnosed at younger ages.(33) Subsequent studies focusing on host genetics, epigenetics, human papillomaviral signatures, and interactions with HIV and other co-factors could shade more light on the development of these histologic variants and how they impact on prognostic outcomes.

Our study findings are unique and are based on the strength of the enrollment protocol jointly developed by investigators from both participating sites in two distinct geographic zones in Nigeria in collaboration with Northwestern University. The histologic diagnoses and tumor grading were further subjected to peer review and quality improvement through a telepathology scanning and video conferencing platform involving the two participating sites and Northwestern pathology core.(18) To the best of our knowledge this is the first of such multidisciplinary team science cervical cancer research project in Nigeria and perhaps in West Africa sub region. However, we acknowledge that despite strenuous follow-up efforts at clinics, cancer treatment centers, and phone contacts with patients and next-of-kin the estimations of the survival outcomes could be limited by incomplete and missing follow-up information on some participants. In addition, though the data on treatment options offered to patients at the participating sites were documented by the gynecologic oncology team, and updated in subsequent follow up visits, the interpretation of the effect size of our survival outcomes is of limited generalizability. We also acknowledge that even though all HIV-positive patients enrolled were taking highly active antiretroviral medications, we did not have enough information on CD4 nor viral load data to confirm immunological recovery and viral load suppression at the time of diagnosis. This information could have helped in subgroup analysis of survival by CD4 and viral load categories. Furthermore, we estimated overall survival and were not able to ascertain disease-free or progression-free survival in the participants.

## Conclusion

Cervical cancer remains a significant public health concern, diagnosed at a relatively young age in women living with HIV, with a significantly lower overall survival probability compared to women without HIV. The trend of presentation and diagnosis of this cancer at advanced stages in women living with HIV could partly explain the differences in overall survival. Strategies that strengthen access to primary prevention, early detection through screening, and early provision of effective treatment should be prioritized for all women-particularly those infected with HIV.

## Author’s Contributions

JM, MAS, RA, AS, LH, RLM, FO, and CJA conceptualized the study and developed the study protocols with support from the pathogenomic laboratory core (JJW, DBG, OS, AF, PA, KB, GI, AA). FM, JA, AM, RA, PA, OS, AF, KB contributed to data collection at the enrollment sites. MK, KB, NK, OO, DN, designed the database and managed project data. MK performed all statistical analysis and interpreted results with JM. JM and MK led the writing of the manuscript with contributions from MAS, RA, BTJ, FM, AM, YZ, EC, NP, JA. All the listed authors contributed to editing of the draft manuscript and approved the final version for submission.

## Data Availability Statement

The relevant data have been presented in the manuscript. Any required data could be provided by the corresponding authors on reasonable request.

## Patients Consent for Publication

Not applicable.

## Ethics approval

The study protocol was reviewed and approved by the Institutional Review Boards (IRB) at UniJos, UniLag, and NU.

## Data Availability

All relevant data are within the manuscript and its supporting information files.

---

**Supplementary file:**
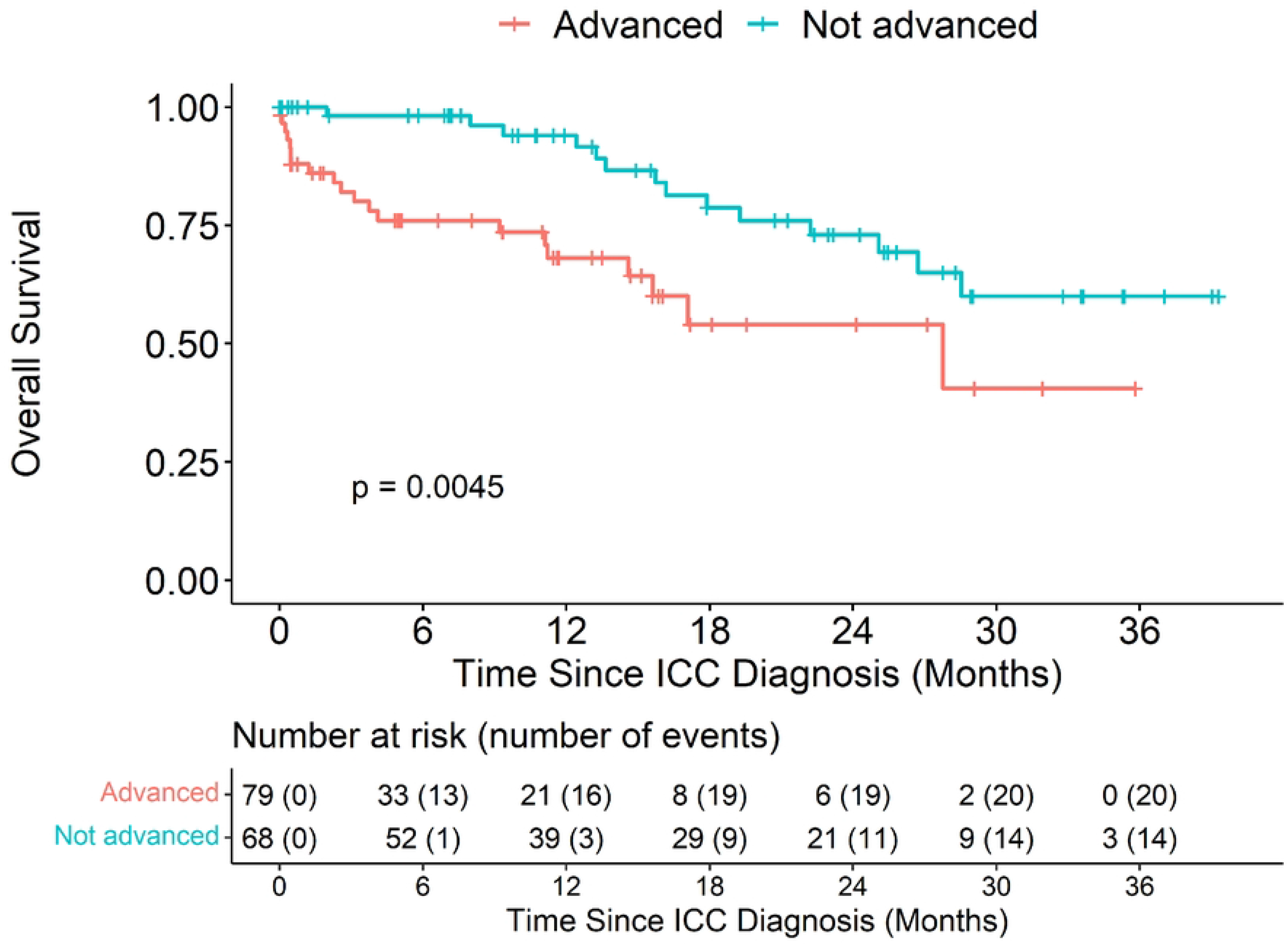
Scanned histopathologic images of cervical cancer with tumor grading

## References

1. Singh D, Vignat J, Lorenzoni V, Eslahi M, Ginsburg O, Lauby-Secretan B, et al. Global estimates of incidence and mortality of cervical cancer in 2020: a baseline analysis of the WHO Global Cervical Cancer Elimination Initiative. Lancet Glob Health. 2023;11(2):e197–e206.

2. Arbyn M, Weiderpass E, Bruni L, de Sanjosé S, Saraiya M, Ferlay J, et al. Estimates of incidence and mortality of cervical cancer in 2018: a worldwide analysis. The Lancet Global Health. 2019.

3. Global Burden of Disease Cancer C, Fitzmaurice C, Allen C, Barber RM, Barregard L, Bhutta ZA, et al. Global, Regional, and National Cancer Incidence, Mortality, Years of Life Lost, Years Lived With Disability, and Disability-Adjusted Life-years for 32 Cancer Groups, 1990 to 2015: A Systematic Analysis for the Global Burden of Disease Study. JAMA Oncol. 2017;3(4):524–48.

4. Ngcobo N, Jaca A, Iwu-Jaja CJ, Mavundza E. Reflection: burden of cervical cancer in Sub-Saharan Africa and progress with HPV vaccination. Curr Opin Immunol. 2021;71:21–6.

5. Stelzle D, Tanaka LF, Lee KK, Ibrahim Khalil A, Baussano I, Shah ASV, et al. Estimates of the global burden of cervical cancer associated with HIV. The Lancet Global Health. 2021;9(2):e161–e9.

6. Eze JN, Emeka-Irem EN, Edegbe FO. A six-year study of the clinical presentation of cervical cancer and the management challenges encountered at a state teaching hospital in southeast Nigeria. Clinical Medicine Insights Oncology. 2013;7:151–8.

7. Anorlu RI. Cervical cancer: the sub-Saharan African perspective. Reproductive Health Matters. 2008;16(32):41–9.

8. Maranga IO, Hampson L, Oliver AW, Gamal A, Gichangi P, Opiyo A, et al. Analysis of factors contributing to the low survival of cervical cancer patients undergoing radiotherapy in Kenya. PLoS One. 2013;8(10):e78411.

9. Musa J, Nankat J, Achenbach CJ, Shambe IH, Taiwo BO, Mandong B, et al. Cervical cancer survival in a resource-limited setting-North Central Nigeria. Infect Agent Cancer. 2016;11:15.

10. Organization WH. Global strategy to accelerate the elimination of cervical cancer as a public health problem. Geneva: World Health organization; 2020.

11. Shah S, Xu M, Mehta P, Zetola NM, Grover S. Differences in Outcomes of Chemoradiation in Women With Invasive Cervical Cancer by Human Immunodeficiency Virus Status: A Systematic Review. Pract Radiat Oncol. 2021;11(1):53–65.

12. Tebeu PM, Ngou-Mve-Ngou JP, Zingue LL, Antaon JSS, Okobalemba Atenguena E, Dohbit JS. The Pattern of Cervical Cancer according to HIV Status in Yaounde, Cameroon. Obstet Gynecol Int. 2021;2021:1999189.

13. Glasmeyer L, McHaro RD, Torres L, Lennemann T, Danstan E, Mwinuka N, et al. Long-term follow-up on HIV infected and non-infected women with cervical cancer from Tanzania: staging, access to cancer-directed therapies and associated survival in a real-life remote setting. BMC Cancer. 2022;22(1):892.

14. Trejo MJ, Lishimpi K, Kalima M, Mwaba CK, Banda L, Chuba A, et al. Effects of HIV status on non-metastatic cervical cancer progression among patients in Lusaka, Zambia. Int J Gynecol Cancer. 2020;30(5):613–8.

15. Nissim O, Dees A, Cooper SL, Patel K, Lazenby GB. Cervical Cancer Among Women With HIV in South Carolina During the Era of Effective Antiretroviral Therapy. J Low Genit Tract Dis. 2022;26(2):109–14.

16. Federal Ministry of Health N. NATIONAL GUIDELINES FOR HIV PREVENTION TREATMENT AND CARE. In: Programme NAaSC, editor. 2020.

17. Matsuo K, Machida H, Mandelbaum RS, Konishi I, Mikami M. Validation of the 2018 FIGO cervical cancer staging system. Gynecol Oncol. 2018.

18. Silas OA, Abdulkareem F, Novo JE, Zheng Y, Nannini DR, Gursel DB, et al. Telepathology in Nigeria for Global Health Collaboration. Ann Glob Health. 2022;88(1):81.

19. Lombe D, M’Ule B C, Msadabwe SC, Chanda E. Gynecological radiation oncology in sub-Saharan Africa: status, problems and considerations for the future. Int J Gynecol Cancer. 2022;32(3):451–6.

20. Semeere A, Freeman E, Wenger M, Glidden D, Bwana M, Kanyesigye M, et al. Updating vital status by tracking in the community among patients with epidemic Kaposi sarcoma who are lost to follow-up in sub-Saharan Africa. BMC Cancer. 2017;17(1):611.

21. Harris PA, Taylor R, Thielke R, Payne J, Gonzalez N, Conde JG. Research electronic data capture (REDCap)--a metadata-driven methodology and workflow process for providing translational research informatics support. J Biomed Inform. 2009;42(2):377–81.

22. Harris PA, Taylor R, Minor BL, Elliott V, Fernandez M, O’Neal L, et al. The REDCap consortium: Building an international community of software platform partners. J Biomed Inform. 2019;95:103208.

23. Team RC. R: A language and environment for statistical computing. R Foundation for Statistical Computing, Vienna, Austria: R Foundation for Statistical Computing, Vienna, Austria; 2022 [Available from: URL https://www.R-project.org/.

24. Odukoya O, Nenrot D, Adelabu H, Katam N, Christian E, Holl J, et al. Application of the research electronic data capture (REDCap) system in a low- and middle income country-experiences, lessons, and challenges. Health Technol (Berl). 2021;11(6):1297–304.

25. Sjoberg DD WK, Curry M, Lavery JA, Larmarange J. Reproducible summary tables with the gtsummary package. The R Journal. 2021;13:570–80.

26. T T. A Package for Survival Analysis in R R package version 3.3-1 ed2022.

27. Kassambara A KM, Biecek P. (2021). _survminer: Drawing Survival Curves using ‘ggplot2’ R package version 0.4.9 ed2021.

28. Dryden-Peterson S, Bvochora-Nsingo M, Suneja G, Efstathiou JA, Grover S, Chiyapo S, et al. HIV Infection and Survival Among Women With Cervical Cancer. Journal of clinical oncology: official journal of the American Society of Clinical Oncology. 2016;34(31):3749–57.

29. Grover S, Mehta P, Wang Q, Bhatia R, Bvochora-Nsingo M, Davey S, et al. Association Between CD4 Count and Chemoradiation Therapy Outcomes Among Cervical Cancer Patients With HIV. J Acquir Immune Defic Syndr. 2020;85(2):201–8.

30. Musa J, Achenbach CJ, Evans CT, Jordan N, Daru PH, Silas O, et al. HIV status, age at cervical Cancer screening and cervical cytology outcomes in an opportunistic screening setting in Nigeria: a 10-year Cross sectional data analysis. Infectious Agents and Cancer. 2019;14(1):43.

31. Gizaw M, Addissie A, Getachew S, Ayele W, Mitiku I, Moelle U, et al. Cervical cancer patients presentation and survival in the only oncology referral hospital, Ethiopia: a retrospective cohort study. Infect Agent Cancer. 2017;12:61.

32. Awolude OA, Oyerinde SO. Invasive Cervical Cancer in Ibadan: Socio-Sexual Characteristics, Clinical Stage at Presentation, Histopathology Distributions and Hiv Status. Afr J Infect Dis. 2019;13(1):32–8.

33. Nicolas-Parraga S, Alemany L, de Sanjose S, Bosch FX, Bravo IG, Ris Hpv TT, et al. Differential HPV16 variant distribution in squamous cell carcinoma, adenocarcinoma and adenosquamous cell carcinoma. Int J Cancer. 2017;140(9):2092–100.

